# Understanding antidepressant change patterns in the UK Biobank

**DOI:** 10.1101/2025.10.13.25337814

**Authors:** Danyang Li, Chris Wai Hang Lo, Cathryn M. Lewis, Evangelos Vassos, Gerome Breen

## Abstract

**Background:** Antidepressants are the most prescribed medications in psychiatry. Medical records provide a valuable opportunity to explore prescribing patterns and to uncover clinical and genetic factors that influence treatment outcomes.

**Methods:** Using longitudinal primary care records from the UK Biobank, we investigated antidepressant change patterns in both depression and non-depression indications. We examined outcomes including the number of antidepressant changes, and discontinuation that is likely due to intolerable side effects or inadequate response. Genetic analyses including heritability estimation, genetic correlation and polygenic score association were performed.

**Results:** A total of 82,633 individuals in the UK Biobank were prescribed at least one of 37 antidepressants. Of these, 28,332 had a depression diagnosis and 24,543 did not. Selective serotonin reuptake inhibitors (SSRIs), including citalopram and fluoxetine, were the most prescribed antidepressants for depression, while tricyclic antidepressants, especially amitriptyline, dominated prescriptions in non-depression indications. Individuals with depression were more likely to stay on antidepressants longer than those without depression, and to follow preferred antidepressant choices that have changed over time. For depression, discontinuation rates for SSRIs were 9% for likely intolerable side effects and 12% for likely nonresponse. Antidepressant change and discontinuation were significantly enriched for psychiatric and somatic conditions, including recurrent depression, anxiety in the depression group, and pain-related conditions in the non-depression group. Genetic analyses identified two novel variants associated with SSRI early discontinuation. Notable genetic overlap was shown between these antidepressant phenotypes and multiple psychiatric and physical traits, and polygenic scores had significant prediction for treatment outcomes.

**Conclusions:** These findings provide a valuable framework for characterising antidepressant change in primary care records and highlight the potential of integrating clinical and genetic data to better understand factors influencing treatment outcomes. Replication in other large-scale medical records will be essential to advance the discovery of genetic variants associated with antidepressant outcomes.

## Introduction

Antidepressants are commonly prescribed for depression, general anxiety, and a number of other indications, such as neuropathic pain, sleep disorders, and migraine (1). Despite their broad and widespread use, treatment with antidepressants often involves a lengthy trial-and-error process. Many individuals discontinue their prescriptions and undergo multiple switches between various antidepressants, largely due to intolerable side effects or inadequate response. Evidence suggested that 20-30% of individuals discontinued antidepressant treatment because of adverse effects (2–4), and only about one-third achieved remission after their first prescribed antidepressant (5). Understanding the factors that contribute to antidepressant discontinuation is essential to improving the health care and outcomes for individuals receiving these medications.

In the United Kingdom, antidepressants are usually prescribed in the primary care, and more than 30 different drugs across several classes have been widely used. For example, tricyclic antidepressants (TCAs) were among the earliest introduced class and were primarily used for depression until the 1990s. Since then, selective serotonin reuptake inhibitors (SSRIs) have become the first-line choice for depression due to their lower overdose risk (6). Some evidence suggests that individuals who discontinued treatment were associated with a history of prior discontinuation, and greater comorbidity burden (2,7). However, most of these findings come from short-term clinical cohorts or retrospective studies. Long-term, real-world prescribing patterns in UK primary care patients of how antidepressant choices shifted over time and the longitudinal nature of treatment discontinuation remain poorly understood.

Beyond clinical indications, antidepressant outcomes can also be influenced by genetic factors. Recent genome-wide association studies (GWASs) from clinical trials have reported 13% of the variance of depression symptom remission during antidepressant treatment could be explained by common genetic variants (5). Other retrospective studies have shown that self-reported side effects can be significantly predicted by polygenic scores (2,8). Electronic health records (EHRs) provide a powerful alternative to clinical trials and retrospective cohorts, with comprehensive longitudinal data and substantially greater statistical power to uncover clinical and genetic influences on antidepressant outcomes (9,10). Therefore, in this study, we used UK Biobank primary care records to investigate antidepressant prescribing patterns in both depression and non-depression indications. We aimed to describe the longitudinal trajectories of antidepressant changes, identify discontinuation patterns that are likely to be caused by side effects or nonresponse, and evaluate clinical and genetic factors associated with these antidepressant phenotypes.

## Methods

### Participants

The UK Biobank recruited approximately 500,000 participants aged 40-79 between 2006 and 2010 through 22 assessment centres across the UK. Health and lifestyle information, as well as measures of hearing and cognitive function, were collected via touchscreen questionnaires and brief verbal interviews. Coded clinical events, such as diagnoses, clinical history, symptoms, prescriptions, and administrative codes were obtained through linkage with primary care records. All participants have informed consent, demonstrating their understanding of the study’s purpose, procedures, potential risks, and benefits before participating. Primary care prescription records were available for ~230,000 participants. Among them, a list of 37 prescribed antidepressants was curated according to the previous study (9) (Supplementary Table 1, Supplementary Methods).

### Antidepressant prescription for individuals with or without depression

In this study, individuals prescribed antidepressants with depression were defined as those who had at least one primary care diagnosis of depressive disorders and no diagnostic codes for psychotic disorders, bipolar disorder, or substance use disorders (9). Prescriptions of amitriptyline and dosulepin were only included if the daily dosage met the therapeutic threshold for depression (≥50 mg for amitriptyline; ≥75 mg for dosulepin). Prescriptions occurring prior to the first recorded depression diagnosis were also excluded. For the individuals prescribed antidepressants without depression, they were included if they had at least one antidepressant prescriptions but without any depression measures in the UK Biobank (Supplementary Methods). Multiple antidepressants that have been prescribed within 5 days were excluded to remove the effect of poly-therapy.

### Antidepressant change and discontinuation

Antidepressant change was defined as a switch from one antidepressant to another in the primary care records. We calculated the total number of different antidepressants prescribed across the overall sample, and within depression and non-depression groups. This measure served as a proxy for complex-to-treat status, representing cases where the indication was not successfully treated. To distinguish the discontinuation due to intolerable side effects or nonresponse, we defined two discontinuation phenotypes among individuals with depression (Supplementary Figure 1):

1. *Early discontinuation likely due to intolerable side effects*: defined as an antidepressant prescribed only once followed by another antidepressant within 40 days, and the same antidepressant was not prescribed before or after within two years. This is to ensure that individuals who discontinued an antidepressant quickly likely due to severe side effects, and the medication was subsequently avoided in primary care practice.
2. *Late discontinuation likely due to nonresponse*: defined as an antidepressant prescribed for at least 42 days, with at least three prescriptions within a six-month episode to ensure an adequate duration of treatment (9), and this drug was switched to another antidepressant within 40 days.

### Hospital inpatient diagnoses of medical conditions

Primary and secondary diagnoses were retrieved from the UK Biobank hospital inpatient records. We used 3-digit ICD-10 codes, covering Chapters I-XIV, Chapter XVIII, and Chapter XXI of the ICD-10. For individuals with multiple records of the same diagnosis, only the first record was used. To extend the follow-up time prior to the ICD-10 introduction in 1995, ICD-9 codes were mapped to the corresponding 3-digit ICD-10 codes using the general equivalence mappings from the Centre for Disease Control (https://ftp.cdc.gov/pub/Health_Statistics/NCHS/Publications/ICD10CM/2018/).

### Statistical analyses

#### Antidepressant change pathways

To explore enriched antidepressant change pathways, we included sequential drug pairs prescribed to >10 individuals. To maximise the statistical power, we focused on individuals initiating the most common antidepressants, which were citalopram and fluoxetine in the depression group, and amitriptyline in the non-depression group. At the first prescription, enrichment of drug pairs was assessed by testing whether these individuals were more likely to switch to specific second antidepressant compared to those starting on other drugs. Logistic regressions were applied with the second antidepressant as the outcome and the first as the exposure, adjusting for sex, age at prescription, prescription year, and data provider (England TTP, England Vision, Wales EMIS/Vision, and Scotland EMIS/Vision, Supplementary Methods). For later stages, we first identified directionality of each drug pair (drugA→drugB vs drugB→drugA) when followed by more than half of individuals. At each number of antidepressant change, we tested whether drugA more frequently preceded drugB relative to other options, using a logistic regression with drugB as the outcome and drugA as the exposure, adjusting for the same covariates described above. Overlapping drug pairs were further combined into longitudinal trajectories of three or more prescriptions (e.g., drugA→drugB and drugB→drugC combined into drugA→drugB→drugC; Supplementary Figure 2). Associations were considered significant at FDR-corrected p<0.05 across all drug pairs within each initial antidepressant treatment.

#### Medical condition enrichment

For each antidepressant change phenotype, we applied a Gamma regression to evaluate the enrichment of medical conditions among individuals with a higher number of antidepressant changes. For each discontinuation phenotype, a logistic regression was used to estimate the risk of medical conditions in individuals who discontinued treatment compared to those who did not (Supplementary Methods). All models were adjusted for sex, data provider, participant birth year, and year of first prescription. Only diagnoses occurring in more than 100 individuals per phenotype were included. Significant diagnoses were considered at p<0.05 after FDR correction across all phenotypes.

### Genetic analyses

#### Genome-wide association

GWASs were conducted using REGENIE v3.1.3 (11) for five phenotypes, including the number of antidepressant changes in the full sample, depression and non-depression groups, as well as early and late discontinuation of SSRI treatment in the depression group, given predominant SSRI use in this population. GWASs were conducted adjusting for batch, genotyping array, the first ten genetic principal components, data provider, birth year, and year of the first prescription. For the X-chromosome, sex was included as an additional covariate. More details of genotyping, imputation and quality control can be found in Supplementary Methods.

#### Heritability estimate, genetic correlation, and polygenic score

Common genetic variant based heritability was estimated using both Genome-wide Complex Trait Analysis (GCTA) and linkage disequilibrium score regression (LDSC) (12,13). Genetic correlations with selected psychiatric and physical traits were computed using LDSC. Polygenic scores (PGSs) were calculated using SBayesR for major depressive disorder (MDD), schizophrenia, bipolar disorder, and attention deficit hyperactivity disorder (ADHD), based on recent large-scale GWASs (14–18). For antidepressant change phenotypes, a Gamma regression was applied to assess the explained variance of PGS associations. For early and late SSRI discontinuation, logistic regressions were applied, with Nagelkerke’s R2 used to evaluate the explained variance. More details can be found in Supplementary Methods.

## Results

### Characteristics of antidepressant prescription

A total of 82,633 UK Biobank participants had at least one recorded antidepressant prescription. The median age at first prescription was 54 years, and 65% were females. The median follow-up time was 9.6 years. Among them, 28,332 individuals had a primary care diagnosis of depression, while 24,543 had no evidence of depression identified in the UK Biobank. Compared to the non-depression group, individuals with a depression diagnosis were more females, had a younger age at the first prescription and had a longer follow-up time (Supplementary Table 2).

In the depression group, the most commonly prescribed antidepressants were citalopram and fluoxetine, whereas amitriptyline was predominant in the non-depression group (Figure 1a, Supplementary Table 3). Around 1990, tricyclic antidepressants such as dosulepin and lofepramine were the most frequent first-line treatments in the depression group. Fluoxetine became dominant between 1990-2000 and citalopram took over in the early 2000s. In the past few years, sertraline emerged as the most common first prescription (Figure 1b). Among those without depression, amitriptyline consistently dominated the first prescription across years. Dosulepin was frequently prescribed in the early 1990s but declined markedly over time. Most antidepressants showed comparable trends at the subsequent prescriptions, though drugs such as venlafaxine, and mirtazapine showed increased proportion compared to the initial treatment in the depression group (Supplementary Figure 3).

**Figure 1.**
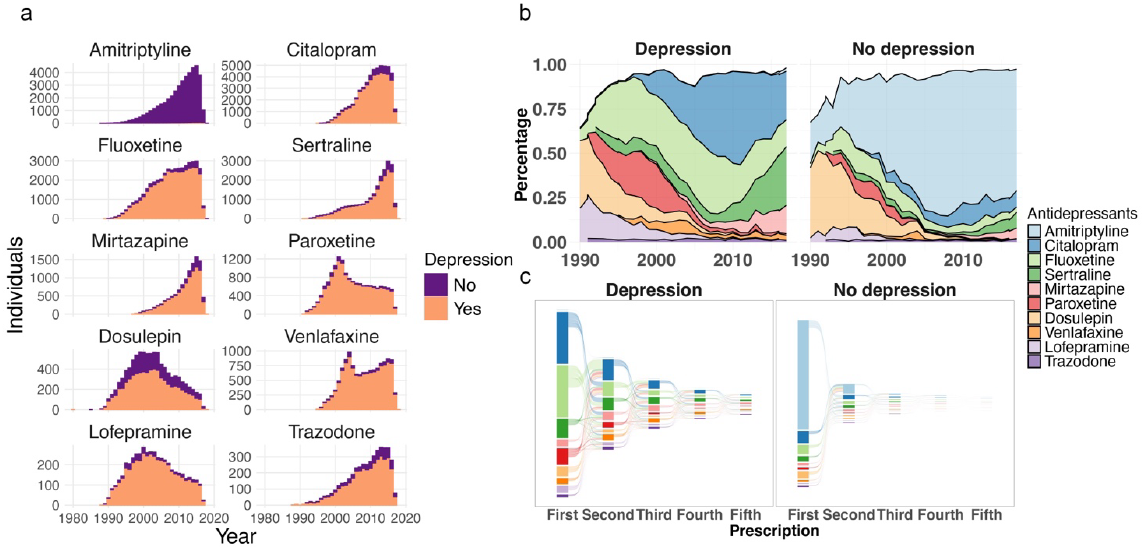
Most prescribed antidepressants in the UK Biobank. a. Number of individuals prescribed antidepressants across years. b. Percentage of the most common ten antidepressants from 1990 to 2018. The blank space represents drugs not listed in the top ten antidepressants. c. Trajectories of the most common ten antidepressants at the first five prescriptions.

The number of antidepressant changes ranged from one to thirteen in the depression group, and from one to nine in the non-depression group (Supplementary Figure 4). Antidepressant changes were more common among individuals with depression, with 47% receiving at least two different antidepressants, compared to 18% in the non-depression group (Supplementary Figure 4). Most individuals with depression started with SSRIs (78%), particularly citalopram (28%), fluoxetine (28%), and sertraline (10%), and 30% continuing SSRIs as the second antidepressant, including citalopram (11%), fluoxetine (8%), followed by 5% of TCAs and 4% of SNRIs. Among individuals without depression, amitriptyline (68%) was overwhelmingly the first-line treatment, with smaller proportions prescribed citalopram (8%) or fluoxetine (6%) (Figure 1c).

### Antidepressant change temporal pathways

We focused on antidepressant change temporal pathways for the most common initial antidepressants, which were citalopram and fluoxetine in the depression group, and amitriptyline in the non-depression group. Across these three initial treatments, we identified 100 drug pairs prescribed to more than ten individuals (Supplementary Table 4). An overview of the drug pair selection and pathway construction can be found in the Supplementary Figure 2. Among individuals starting from citalopram, the most frequent subsequent prescriptions were sertraline (27%), fluoxetine (24%), mirtazapine (17%), and escitalopram (5%). Drugs like sertraline, mirtazapine, and venlafaxine were most often prescribed as the third prescription (Supplementary Figure 5). For those starting with fluoxetine, citalopram (37%), sertraline (16%), venlafaxine (9%), and paroxetine (9%) were more likely to be prescribed as the second antidepressant. Switching from fluoxetine to citalopram was frequently followed by sertraline or mirtazapine, while switching to dosulepin, escitalopram, paroxetine, or venlafaxine often led back to citalopram, which in turn was followed by sertraline or mirtazapine (Supplementary Figure 6). In the non-depression group, individuals beginning with amitriptyline were more likely to switch to citalopram (22%), nortriptyline (16%), fluoxetine (13%), or duloxetine (9%), and citalopram was commonly followed by mirtazapine or sertraline (Supplementary Figure 7). All significant drug pairs stratified by the year of prescription could be found in Supplementary Figure 8.

### Antidepressant discontinuation likely due to intolerable side effects or nonresponse in individuals with depression

We defined two types of antidepressant discontinuation, where early discontinuation reflected treatment cessation likely due to intolerable side effects, and late discontinuation indicated cessation more likely attributable to lack of response (Supplementary Figure 1). In the depression group, 9% of individuals experienced at least one early discontinuation, while 12% had at least one late discontinuation. In each drug class, TCAs showed the highest rate of early discontinuation (7.3%), followed by SSRIs (5.9%) and SNRIs (5.2%) (χ^2^=19.75, p=5.14×10^−5^). In contrast, late discontinuation was most frequent among SSRIs (9.9%) compared to SNRIs (9.2%) and TCAs (8.8%) (χ^2^=6.25, p=0.044, Supplementary Table 5). Both discontinuation rates steadily increased across the first seven antidepressants (Supplementary Figure 9). Compared to SSRIs and SNRIs, TCAs consistently showed higher early discontinuation rates. For the late discontinuation, the three drug classes had comparable rates during the first three antidepressants, but from the fourth antidepressant onward, SSRIs showed a higher late discontinuation rate than SNRIs or TCAs (Supplementary Figure 10).

### Enrichment of medical conditions in antidepressant change and discontinuation

From hospital inpatient diagnoses, we identified 207 medical conditions in the depression group and 187 in the non-depression group with more than 100 individuals each. Of these, 158 and 146 conditions showed significant associations with antidepressant change in depression and non-depression groups, respectively. In the depression group, frequent antidepressant change was most strongly enriched for recurrent depression, depressive episode, anxiety disorders, other brain disorders, pain syndromes, and speech disturbances. In the non-depression group, enrichment was observed for anxiety, pain, hemiplegia, and polyneuropathies (Figure 2, Supplementary Table 6). For SSRI discontinuation phenotypes, early discontinuation was associated with recurrent depression, other brain disorders, chronic pain (R30, R52), and somatic conditions including cardiac arrhythmias, paroxysmal tachycardia, intestinal malabsorption, and irritable bowel syndrome. Late discontinuation was most strongly associated with recurrent depression, depressive episode, anxiety, other brain disorders, speech disturbances, pain-related conditions (e.g., arthrosis of the first carpometacarpal joint (M18), pain (R52)), and somatic disorders such as obesity and respiratory failure (Supplementary Table 6).

**Figure 2.**
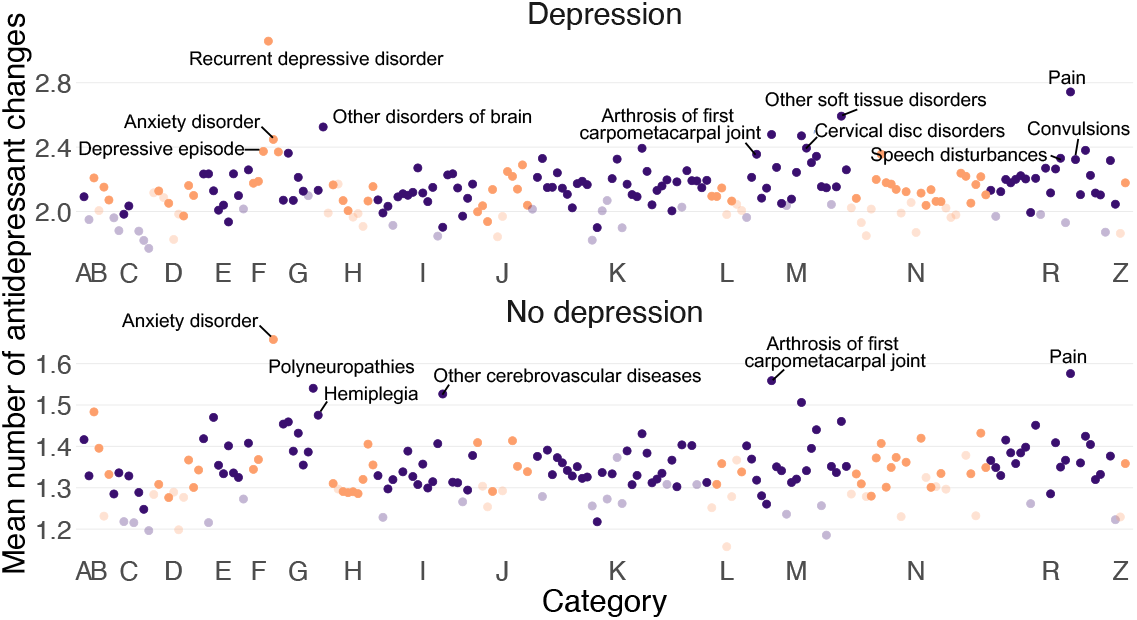
Mean number of antidepressant changes of each medical condition among individuals with or without depression. The X axis shows the medical condition categories of ICD-10 codes A-N, R and Z. The Y axis shows the mean number of antidepressant changes of each medical condition in individuals with or without depression. Each dot represents a hospital inpatient diagnosis. Diagnoses within ICD-10 categories A, C, E, G, I, K, M, and R are shown in purple, and those in categories B, D, F, H, J, L, N, and Z are shown in orange. The dark colour indicates statistically significant diagnoses, and the light colour shows nonsignificant ones. Details of the effect size, 95% confidence intervals, and sample size are listed in the Supplementary Table 6.

### SNP-based heritability and genetic correlation

To further investigate genetic factors influencing antidepressant change and discontinuation, we performed genome-wide association for five antidepressant phenotypes, including the number of antidepressant changes in the full population, depression, and non-depression groups, as well as SSRI early and late discontinuation in the depression group (Table 1). Two variants showed genome-wide significance for SSRI early discontinuation in the depression group (rs184183447: beta=1.29, p=2.90×10^−10^, minor allele frequency=0.011; rs142966989: beta=1.29, p=7.11×10^−10^, minor allele frequency=0.011). They were located nearest to the *FLT1* gene. No significant variants were identified for the other phenotypes (Supplementary Figure 11-15). LDSC intercepts were about 1 in all these GWASs, suggesting no confounding factors (Supplementary Figure 11-15).

**Table 1.** GWAS sample size of number of antidepressant changes and discontinuation phenotypes.

| Phenotypes | N | Have discontinuation | No discontinuation |
| --- | --- | --- | --- |
| Number of antidepressant changes |  |  |  |
| All | 75,421 |  |  |
| Depression | 28,332 |  |  |
| Non-depression | 24,378 |  |  |
| SSRI discontinuation in the depression group |  |  |  |
| Early | 19,148 | 1,342 | 17,806 |
| Late | 17,256 | 2,315 | 14,941 |

SNP-based heritability estimates for the number of antidepressant changes were comparable across groups, e.g., GCTA estimates were 2.6% (SE=0.0053, p=1.89×10^−7^), 2.5% (SE=0.014, p=0.034), and 2.8% (SE=0.017, p=0.040) in the full population, depression, and non-depression groups, respectively. Discontinuation phenotypes showed higher heritability, with GCTA estimates of 9.1% for early discontinuation and 4.6% for late discontinuation, although they did not reach statistical significance (early discontinuation: SE=0.067, p=0.084, late discontinuation: SE=0.052, p=0.19, Supplementary Figure 16). Genetic correlation between antidepressant phenotypes can be found in Supplementary Table 7, and none were significantly different from zero.

We next tested the genetic correlation of antidepressant phenotypes with various psychiatric and physical traits. The number of antidepressant changes in the full population was most strongly correlated with anxiety and major depressive disorder (MDD) (r_g_=0.81-0.83, Figure 3). Substantial genetic correlation was found with irritable bowel syndrome (IBS), chronic pain, insomnia, ADHD, neuroticism, arrhythmia, and dizziness (r_g_=0.49-0.67). Moderate correlations were identified with tachycardia, bipolar disorder, migraine, smoking and schizophrenia (r_g_=0.27-0.33). Negative correlations were observed with educational attainment (r_g_=−0.44). Compared to the non-depression group, antidepressant change in the depression group had stronger genetic correlations with most psychiatric and neurological traits, including anxiety, MDD, insomnia, and migraine (r_g_=0.47-0.72), while the non-depression group was more strongly correlated with chronic pain (r_g_=0.61), although the estimate did not reach statistical significance. For discontinuation phenotypes, early discontinuation was significantly correlated with educational attainment (r_g_=−0.17), and had the strongest correlation with arrhythmia (r_g_=0.21), though the correlation was not significant. Late discontinuation showed the highest correlation with ADHD (r_g_=0.29), followed by MDD (r_g_=0.23), insomnia (r_g_=0.20), dizziness (r_g_=0.18), smoking (r_g_=0.16), and educational attainment (r_g_=−0.13).

### Polygenic scores

We calculated PGSs for MDD, schizophrenia, bipolar disorder, and ADHD. The MDD PGS explained 0.22% (adjusted p=1.05×10^−107^) of the variance in the number of antidepressant changes in the full population, 0.14% (adjusted p=2.50×10^−22^) in the depression group, and 0.02% (adjusted p=4.30×10^−6^) in the non-depression group (Figure 4). Significant associations were also observed for the ADHD PGS across all three groups, and for the bipolar disorder and schizophrenia PGSs in the full population and depression group (Figure 4, Supplementary Table 8). For SSRI early discontinuation, the MDD PGS explained 0.03% (adjusted p=0.49) of the variance and ADHD PGS accounted for 0.05% (adjusted p=0.34). For SSRI late discontinuation, PGS for ADHD showed the strongest association with the explained variance of 0.64% (adjusted p=5.07×10^−7^), followed by PGS for MDD (liability r2=0.35%, adjusted p=2.33×10^−5^) and bipolar disorder (liability r2=0.23%, adjusted p=0.0036).

**Figure 3.**
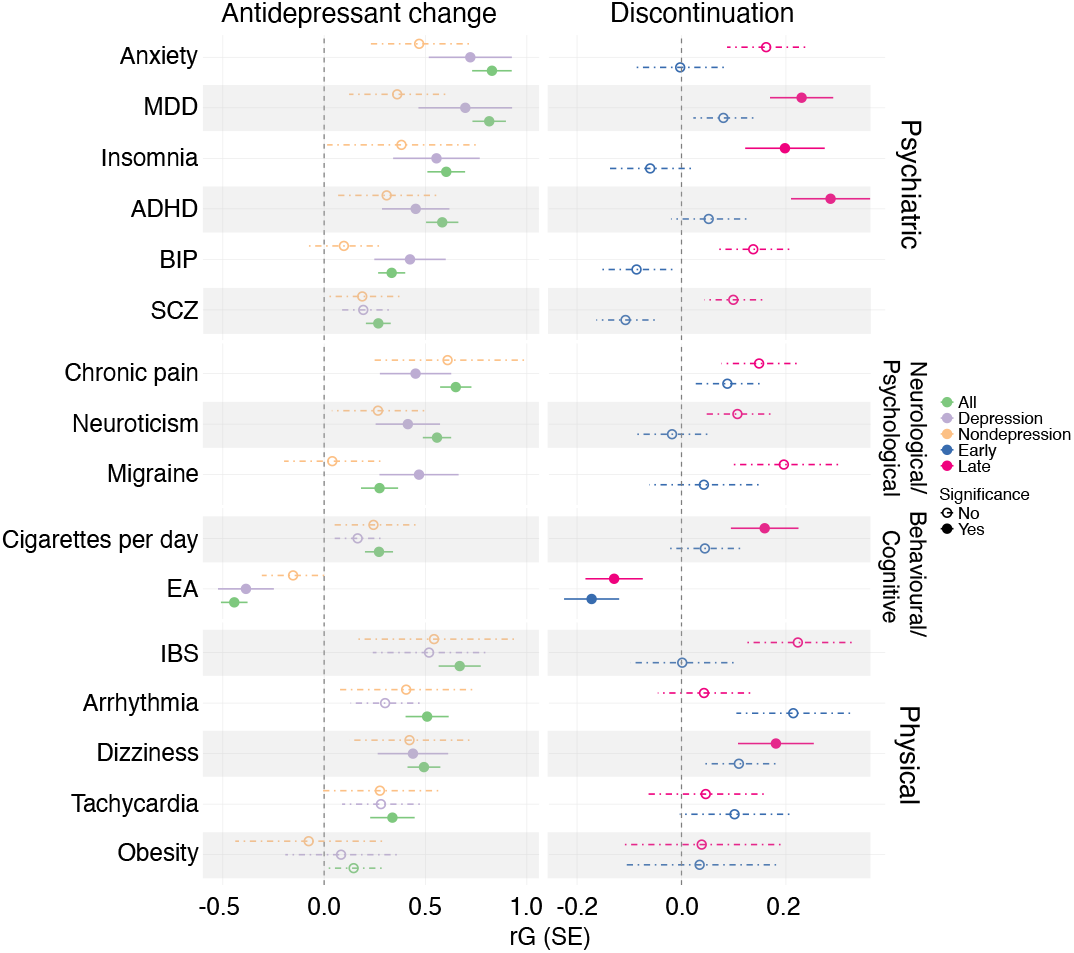
Genetic correlation of antidepressant change and discontinuation with psychiatric and physical traits. MDD: major depressive disorder; ADHD: attention deficit hyperactivity disorder; BIP: bipolar disorder; SCZ: schizophrenia; EA: educational attainment; IBS: irritable bowel syndrome

**Figure 4.**
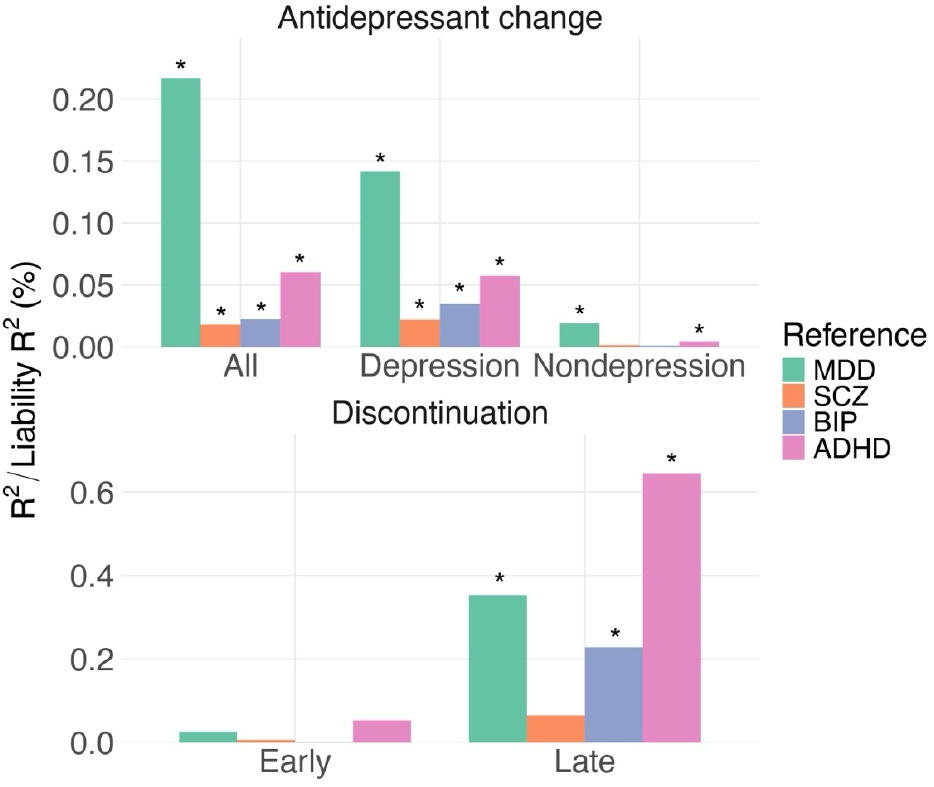
Explained variance of polygenic score for antidepressant change and discontinuation phenotypes. MDD: major depressive disorder; SCZ: schizophrenia; BIP: bipolar disorder; ADHD: attention deficit hyperactivity disorder. *: adjusted p-value < 0.05

## Discussion

Our study is the first to characterise the longitudinal landscape of antidepressant change and discontinuation patterns for both depression and non-depression indications in the UK Biobank. Using primary care records, we found that individuals with depression were most frequently prescribed SSRIs, such as fluoxetine, citalopram, and sertraline, whereas TCAs, particularly amitriptyline, were most prescribed among those without depression. Over the past 30 years, the preferred antidepressant choices have shifted markedly, showing evolving clinical recommendations in the UK primary care. We also identified two discontinuation phenotypes likely reflecting intolerable side effects or inadequate response. All these phenotypes were enriched for a range of psychiatric and somatic conditions. Genetic analyses revealed substantial overlap between antidepressant phenotypes and multiple psychiatric and physical traits, and polygenic scores could explain a notable proportion of outcome variance.

We observed distinct patterns of antidepressant prescribing between the depression and non-depression groups. Consistent with previous reports, SSRI prescription for depression, including fluoxetine and citalopram rose rapidly from the early 1990s, followed by a marked increase of sertraline after 2009, due to its favourable balance between efficacy and tolerability (19–21). Among TCAs, use of dosulepin and lofepramine declined steadily after 1990, while low-dose amitriptyline increased substantially, covering a range of indications, such as chronic pain, sleep disorders, anxiety, migraine, psychological distress, and irritable bowel syndrome (19,20,22).

We observed that individuals with a depression diagnosis were more likely to switch to another antidepressant and follow longer switching pathways compared to those without a depression diagnosis. However, these pathways were complex and varied over time. For example, individuals starting on fluoxetine could switch to other SSRIs, such as citalopram or sertraline, or to TCAs like lofepramine and dosulepin (23). However, switches to TCAs mainly occurred before the early 2000s, and changing to SSRIs was preferred in later years, in line with updated clinical guidelines recommending the use of an alternative SSRI before moving to less tolerated drugs (21). The discontinuation rates likely due to intolerable side effects or nonresponse were both lower than those reported in self-reported cohorts and clinical trials (2–5,24). This difference is likely due to our stringent phenotype definitions using treatment continuation and timing of drug switches (9), in the context of the absence of clinically measured treatment response and underreported side effect symptoms in the structured fields of primary care records (25). TCAs showed the highest early discontinuation rates, consistent with their known tolerability issues (2,26). In contrast, late discontinuation rates were initially similar across drug classes but diverged after the third antidepressant, with SSRIs showing higher rates. As primary care physicians may consider switching to TCAs after SSRIs nonresponse, and our definition only included monotherapy switches excluding drug augmentation and other treatment strategies such as electroconvulsive therapy, these results need to be interpreted with caution.

Further analyses of hospital inpatient diagnoses showed that frequent antidepressant change was associated with anxiety, pain conditions, and somatic diagnoses that can cause pain-related symptoms, such as hemiplegia, migraine, and osteoarthritis. In the depression group, additional associations included recurrent depression, speech disturbance, and brain disorders (e.g. encephalopathy). In addition to psychiatric disorders, early discontinuation was linked to cardiac disorders such as arrhythmias, and gastrointestinal issues including intestinal malabsorption, and irritable bowel syndrome. These somatic diagnoses have commonly been reported as concerning side effects for SSRIs (27,28). Late discontinuation was associated with obesity, in line with previous studies showing that a higher BMI reduced effectiveness and increased risk of treatment resistance (29,30).

For the variants associated with SSRI early discontinuation (rs184183447 chr13:28515228, rs142966989 chr13:28496354), the nearest gene is *FLT1* (chr13:28300346-28495128), encoding the Vascular Endothelial Growth Factor Receptor 1, which is also known as *VEGFR1*. The VEGF signalling pathway including FLT1 is important in neurogenesis and synaptogenesis, and it has been reported to be associated with the severity of depressive symptoms (31). Clinical and GWAS studies also revealed the role of *FLT1* on suicide attempt (31–33), which was one of the severe side effects caused by SSRIs. Nevertheless, the strength of these GWAS signals was modest, and replication in larger cohorts is needed.

The number of antidepressant changes reflected the overall complexity of treatment management, but it carried heterogeneous causes which could be side effect, nonresponse, or other treatment factors. In contrast, early and late discontinuation phenotypes aimed to maximise the specific cause of treatment outcomes. Consistent with this, SNP-based heritability especially for the early discontinuation of SSRIs reached 9%, higher than the number of antidepressant changes around 2%. This estimate also exceeded the earlier study of 4% capturing SSRIs switching from primary care records (23). Although the heritability for discontinuation phenotypes were not significantly different from zero, probably due to limited sample size, it highlighted the potential value of future meta-analyses to integrate EHR data across multiple resources to achieve more reliable findings (34,35).

Genetic correlations showed a shared genetic predisposition between antidepressant change and a range of psychiatric disorders, such as MDD, neuroticism, ADHD, consistent with previous findings of treatment resistant depression (9). As expected, the depression group showed higher correlations with most psychiatric and neurological traits, including anxiety, MDD, neuroticism, insomnia, bipolar disorder, migraine, while the non-depression group tend to have stronger correlations with chronic pain and physical conditions including IBS. For the discontinuation phenotypes, although most traits were not significantly correlated with early discontinuation, we observed the highest genetic correlation between early discontinuation and arrhythmia, suggesting a shared genetic liability to traits related to specific side effects, especially cardiac symptoms. As in prior studies of self-reported nonresponse and treatment resistant depression (9,24), we identified a strong genetic correlation and PGS association between late discontinuation and ADHD. Notably it has been reported that in primary care, fewer than 20% of adult ADHD cases were detected (36) and studies show that ADHD could increase the risk of antidepressant treatment resistance (37,38). One possible explanation was emotional dysregulation, which was worsened when ADHD co-occurred with major depression and further increased the risk of treatment resistance (38). These findings underscore the importance of effective and reliable ADHD assessment, particularly for individuals showing poor antidepressant response.

Some limitations should be noted in our study. The primary care EHR data covered only about half of UK Biobank participants, limiting overlap with other subset of samples. Available prescription records were primarily from the 1990s onward, so treatment histories for older antidepressants, such as TCAs, were incomplete. Although there were no clinical measures of response such as depression symptom before and after the treatment, and most side effects were likely underreported in the structured EHR data (25), the early and late discontinuation we measured using treatment continuation and antidepressant switching has been reported in treatment resistant depression (9,39) and our genetic findings also showed good concordance with self-reported outcomes (24). Given the variety of available depression treatments, future work should incorporate other treatment options such as psychotherapy, electroconvulsive therapy, and augmentation therapies (lithium, atypical antipsychotics) to fully assess the antidepressant discontinuation patterns in the context of broader treatment. While our genetic analyses had sufficient power to detect heritability for the number of antidepressant changes, they were underpowered for the discontinuation phenotypes. Larger sample sizes combining other EHR resources will be necessary to validate our findings. Lastly, given that UK Biobank participants were primarily of European ancestry from the UK, further studies are required using primary care records with diverse population ancestries.

In summary, our study is the first to investigate the longitudinal antidepressant changes using primary care records from the UK Biobank. We identified distinct prescription patterns between individuals with and without depression. We defined the number of antidepressant changes and discontinuation likely due to side effects and nonresponse, and our findings showed that individuals with frequent antidepressant changes or discontinuation had significantly different genetic and clinical characteristics compared to those with fewer antidepressant changes or without discontinuation due to side effects or nonresponse. Our study provides a valuable framework for assessing antidepressant prescribing patterns in EHR data and highlights the potential of integrating clinical and genetic information to better understand factors influencing treatment outcomes, ultimately supporting more personalised approaches to antidepressant treatment.

## Acknowledgements

This research has been conducted using the UK Biobank Resource under Application Number 82087. We are grateful to the UK Biobank participants and staff for their valuable contributions. Analyses were performed on the CREATE High Performance Computer at King’s College London and UK Biobank Research Analysis Platform. We especially thank Dr Chiara Fabbri for kindly providing valuable support on the antidepressant identification and depression diagnosis in the primary care records.

## Authors’ contributions

Conception and design: DL. Supervision: CML, EV, GB. Data analysis: DL, CWHL. Funding: CML, GB. First draft: DL. Draft revision and approval: all authors.

## Data and code availability

UKB data are publicly available to bona fide researchers upon application at http://www.ukbiobank.ac.uk/using-the-resource/. All software and analytical methods used in this study are publicly available. Analysis code will be accessible on github shortly.

