## Supplementary Material for "Understanding antidepressant change patterns in the UK Biobank"

*Supplementary Figures* ... *3*

Supplementary Figure 1. Definition of discontinuation phenotypes ... 3

Supplementary Figure 2. Overview of antidepressant change pathway identification in the UK Biobank ... 4

Supplementary Figure 3. Proportion of the most ten common antidepressants prescribed at the second and later prescriptions ... 5

Supplementary Figure 4. Total number of antidepressant changes in depression and non-depression groups ... 6

Supplementary Figure 5. Antidepressant change pathways starting from citalopram in the depression group ... 7

Supplementary Figure 6. Antidepressant change pathways starting from fluoxetine in the depression group ... 8

Supplementary Figure 7. Antidepressant change pathways starting from amitriptyline in the non-depression group ... 9

Supplementary Figure 8. Percentage of significant drug pairs changed across years ... 10

Supplementary Figure 9. Early and late discontinuation rates at the first ten prescribed antidepressants ... 11

Supplementary Figure 10. Early and late discontinuation rates of each antidepressant class across number of different antidepressants prescribed ... 12

Supplementary Figure 11. GWAS association of the number of antidepressant changes in all population ... 13

Supplementary Figure 12. GWAS association of the number of antidepressant changes in the depression group ... 14

Supplementary Figure 13. GWAS association of the number of antidepressant changes in the non-depression group ... 15

Supplementary Figure 14. GWAS association of SSRI early discontinuation ... 16

Supplementary Figure 15. GWAS association of SSRI late discontinuation ... 17

Supplementary Figure 16. SNP heritability (SNP-h2) estimated by GCTA and LDSC ... 18

*Supplementary Tables* ... *19*

Supplementary Table 1. Prescribed antidepressants in the UK Biobank ... 19

Supplementary Table 2. Characteristics of individuals with antidepressant prescription in depression and non-depression groups. ... 20

Supplementary Table 3. The top ten most prescribed antidepressants in the UK Biobank ... 21

Supplementary Table 5. Early and late discontinuation rates across antidepressant classes in individuals with depression ... 22

Supplementary Table 7. Genetic correlation among antidepressant change and discontinuation phenotypes ........ 23

*Supplementary Methods* ........ *24*

### Supplementary Figures

#### Supplementary Figure 1. Definition of discontinuation phenotypes

Early discontinuation likely due to intolerable side effects:

1. An antidepressant prescribed only once followed by another antidepressant within 40 days
2. This antidepressant is not prescribed before or after within 2 years

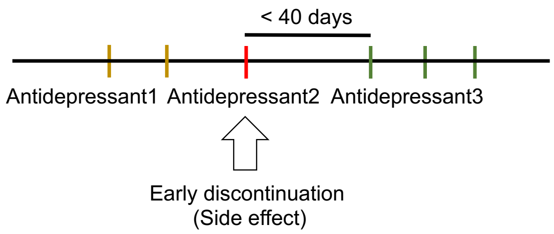


Late discontinuation likely due to nonresponse:

1. An antidepressant prescribed at least 6 weeks in a prescription episode
2. This antidepressant was prescribed more than 3 times in this episode
3. This drug was switched to another antidepressant within 40 days

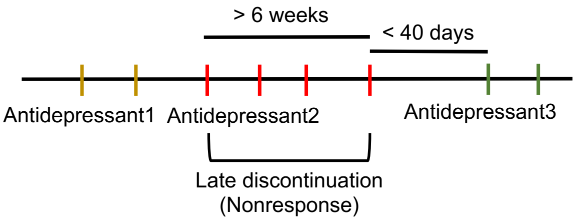

Supplementary Figure 2. Overview of antidepressant change pathway identification in the UK Biobank

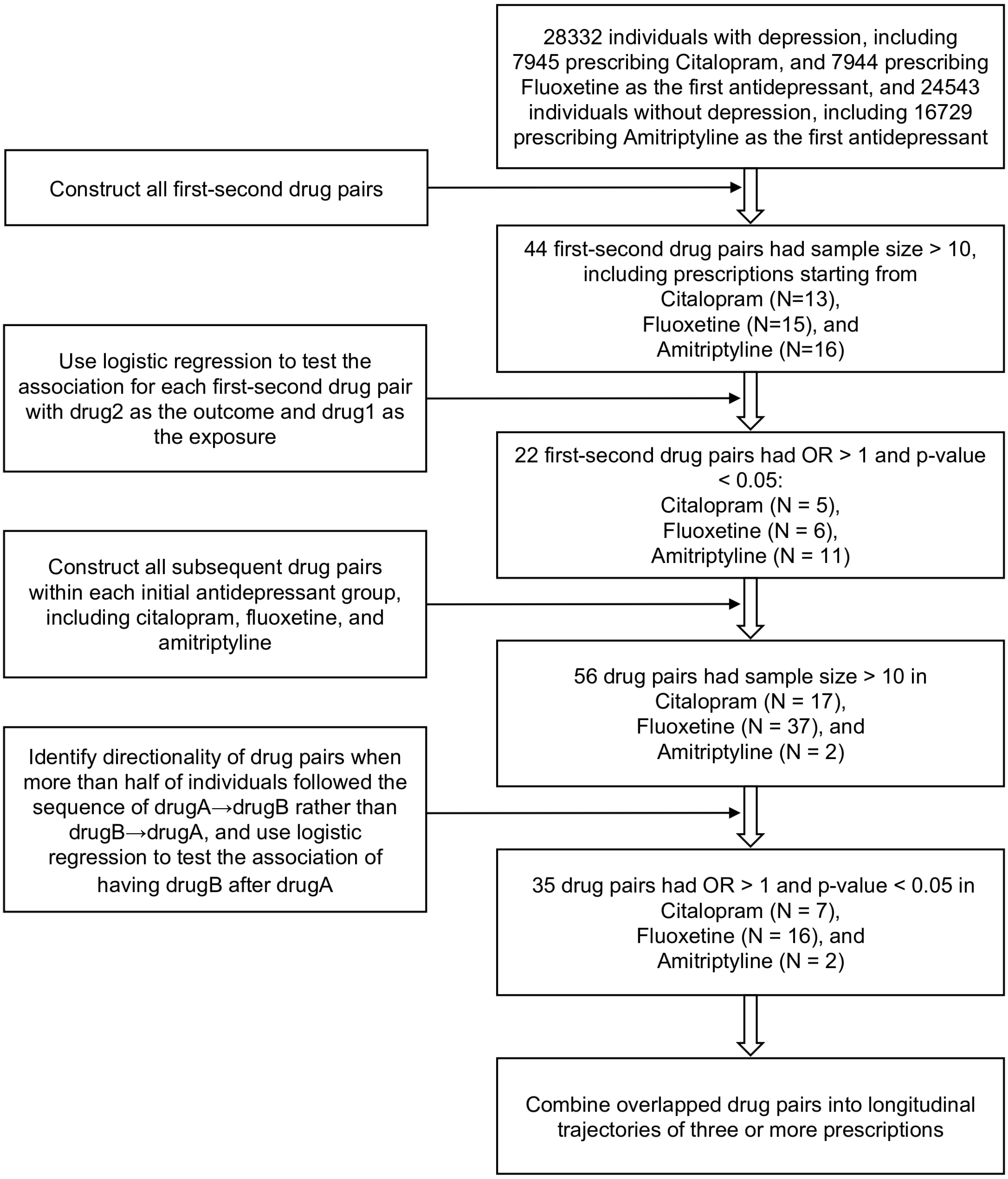

Supplementary Figure 3. Proportion of the most ten common antidepressants prescribed at the second and later prescriptions

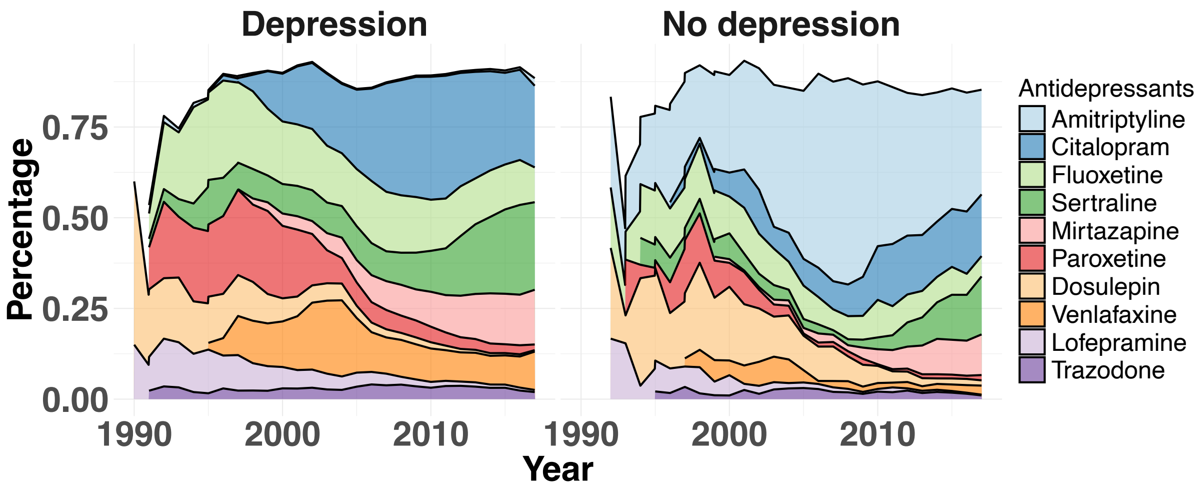

Supplementary Figure 4. Total number of antidepressant changes in depression and non-depression groups

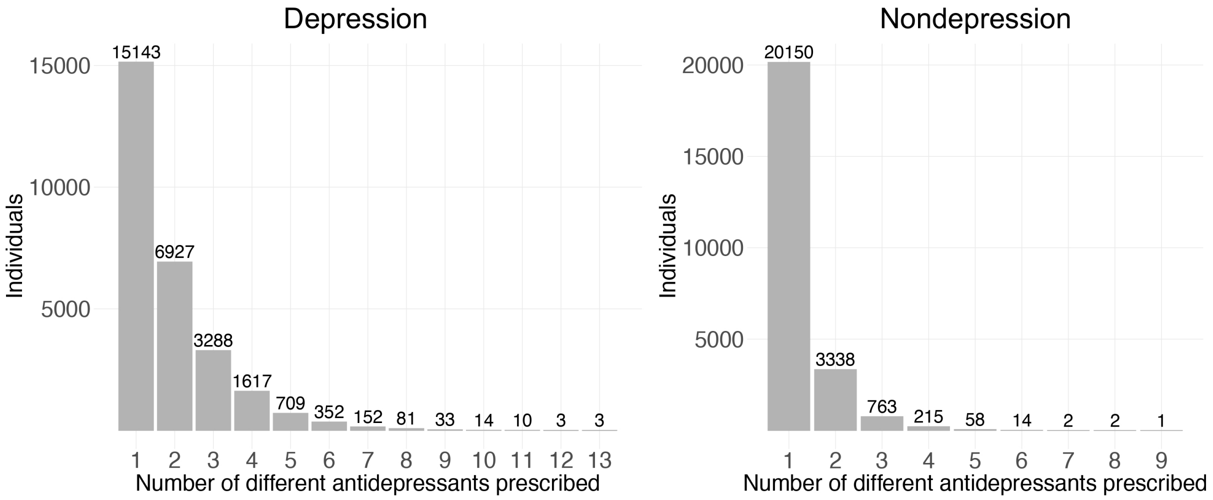

Supplementary Figure 5. Antidepressant change pathways starting from citalopram in the depression group

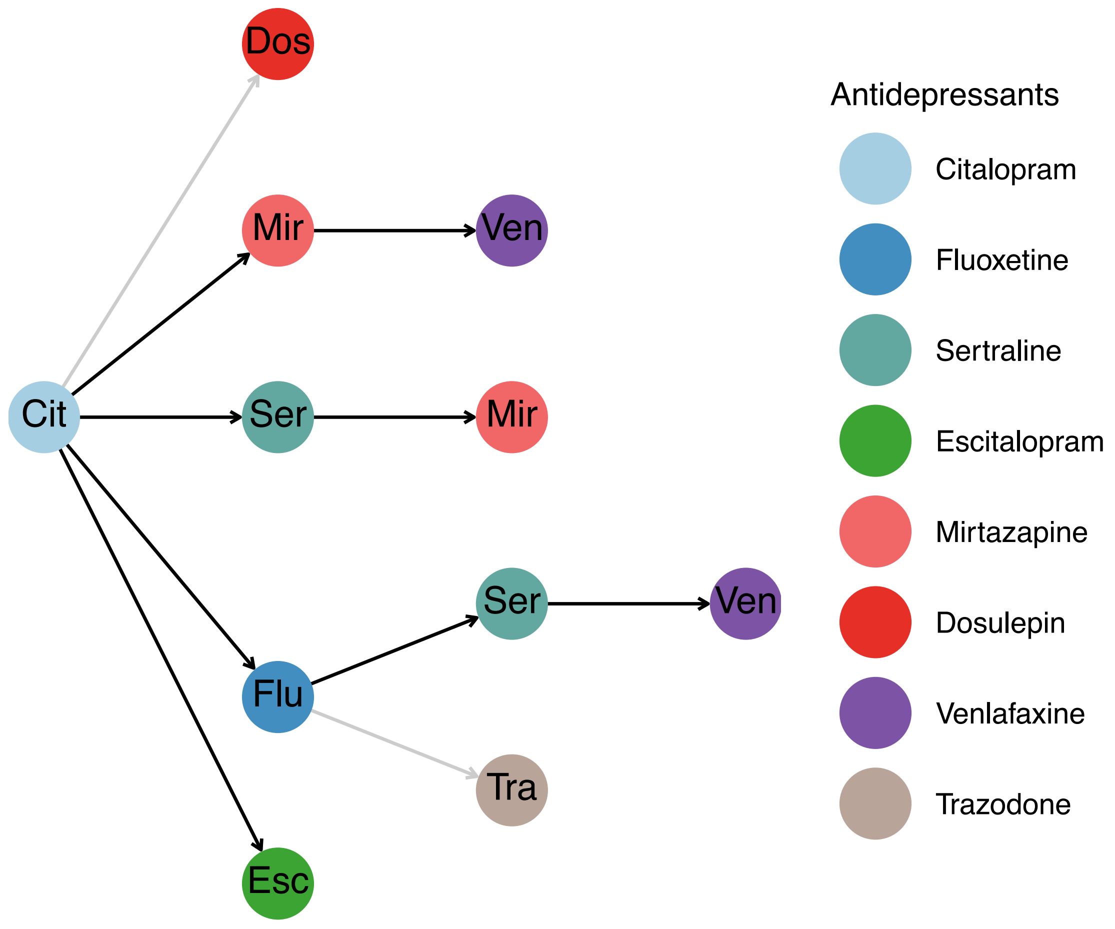


All drug pairs shown in the figure had the sample size > 10, OR > 1, and association results with the nominal significant level of P < 0.05, with the grey line showing unadjusted P < 0.05, and the black line of FDR adjusted P < 0.05

Supplementary Figure 6. Antidepressant change pathways starting from fluoxetine in the depression group

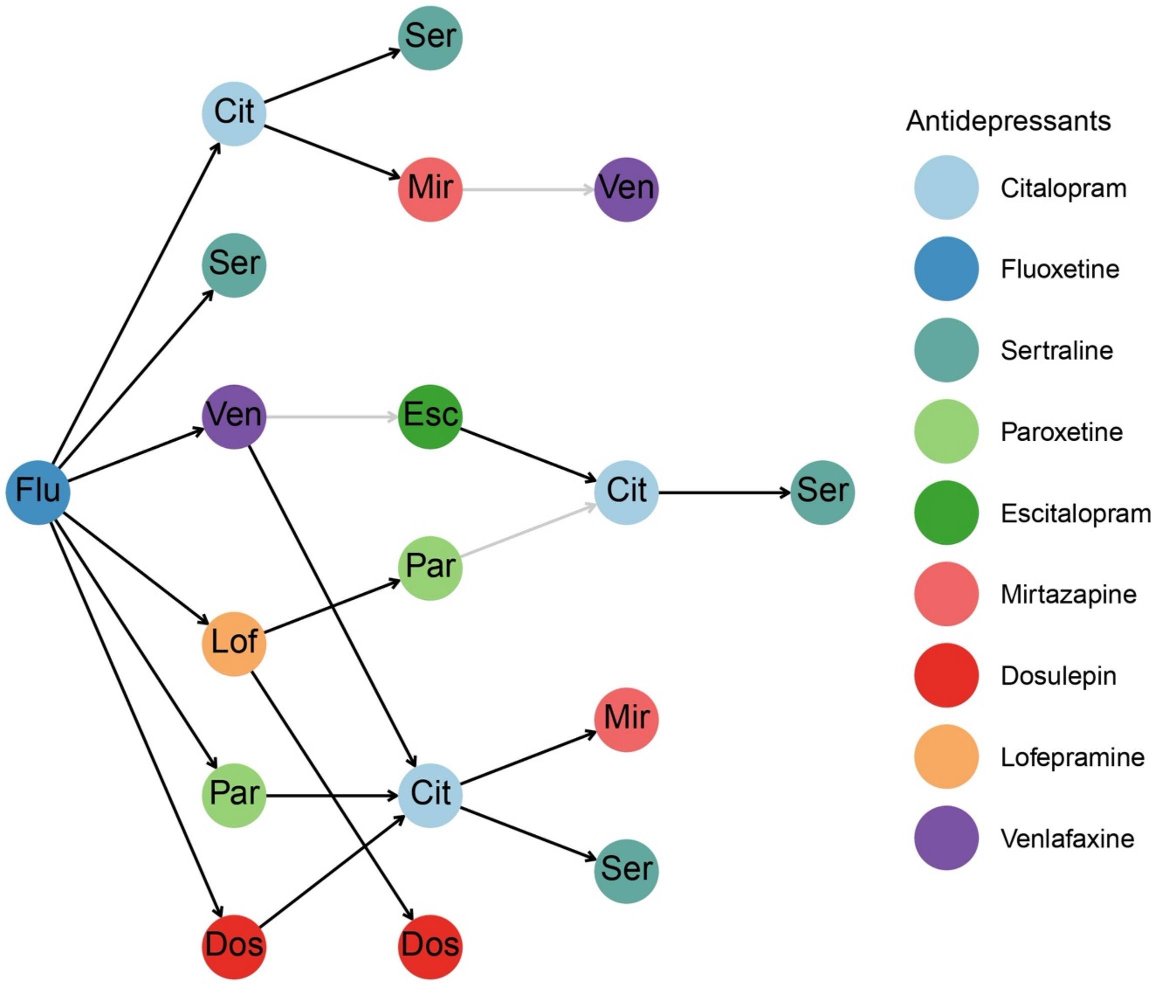


All drug pairs shown in the figure had the sample size > 10, OR > 1, and association results with the nominal significant level of P < 0.05, with the grey line showing unadjusted P < 0.05, and the black line of FDR adjusted P < 0.05

Supplementary Figure 7. Antidepressant change pathways starting from amitriptyline in the non-depression group

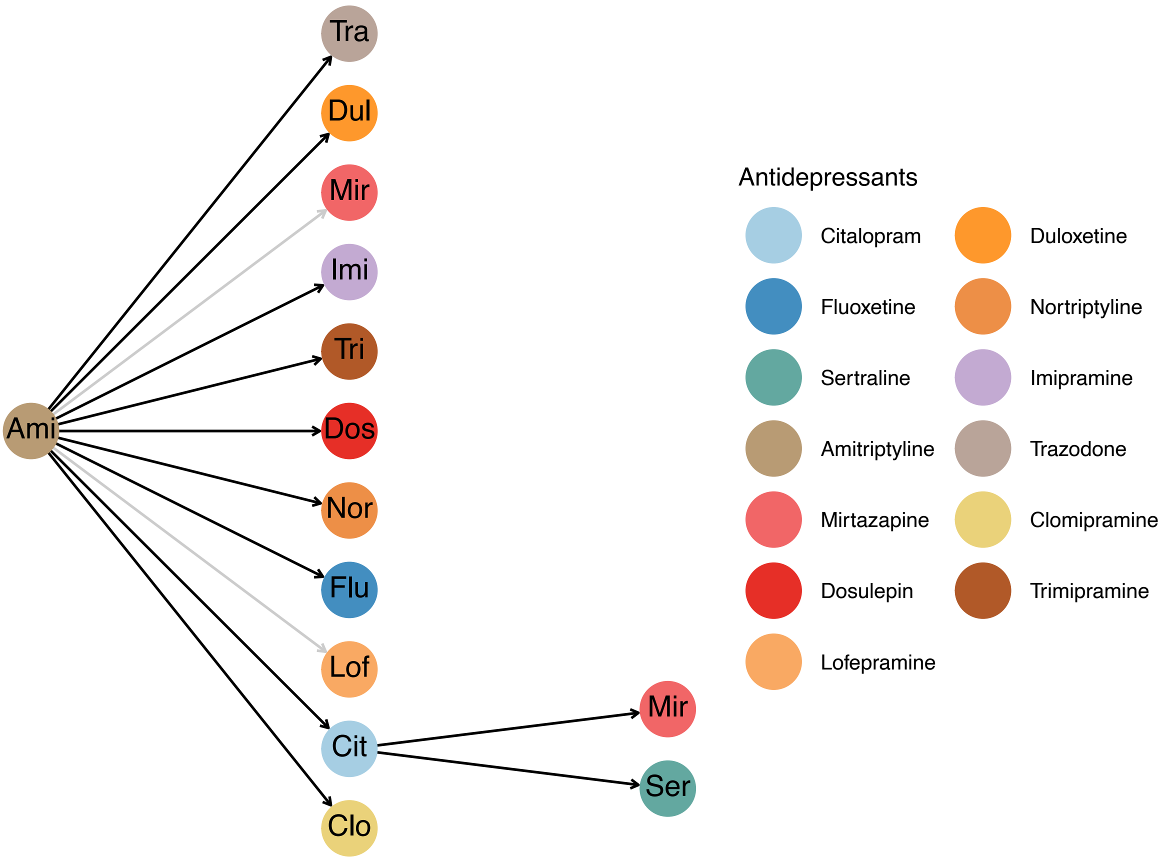


All drug pairs shown in the figure had the sample size > 10, OR > 1, and association results with the nominal significant level of P < 0.05, with the grey line showing unadjusted P < 0.05, and the black line of FDR adjusted P < 0.05

Supplementary Figure 8. Percentage of significant drug pairs changed across years

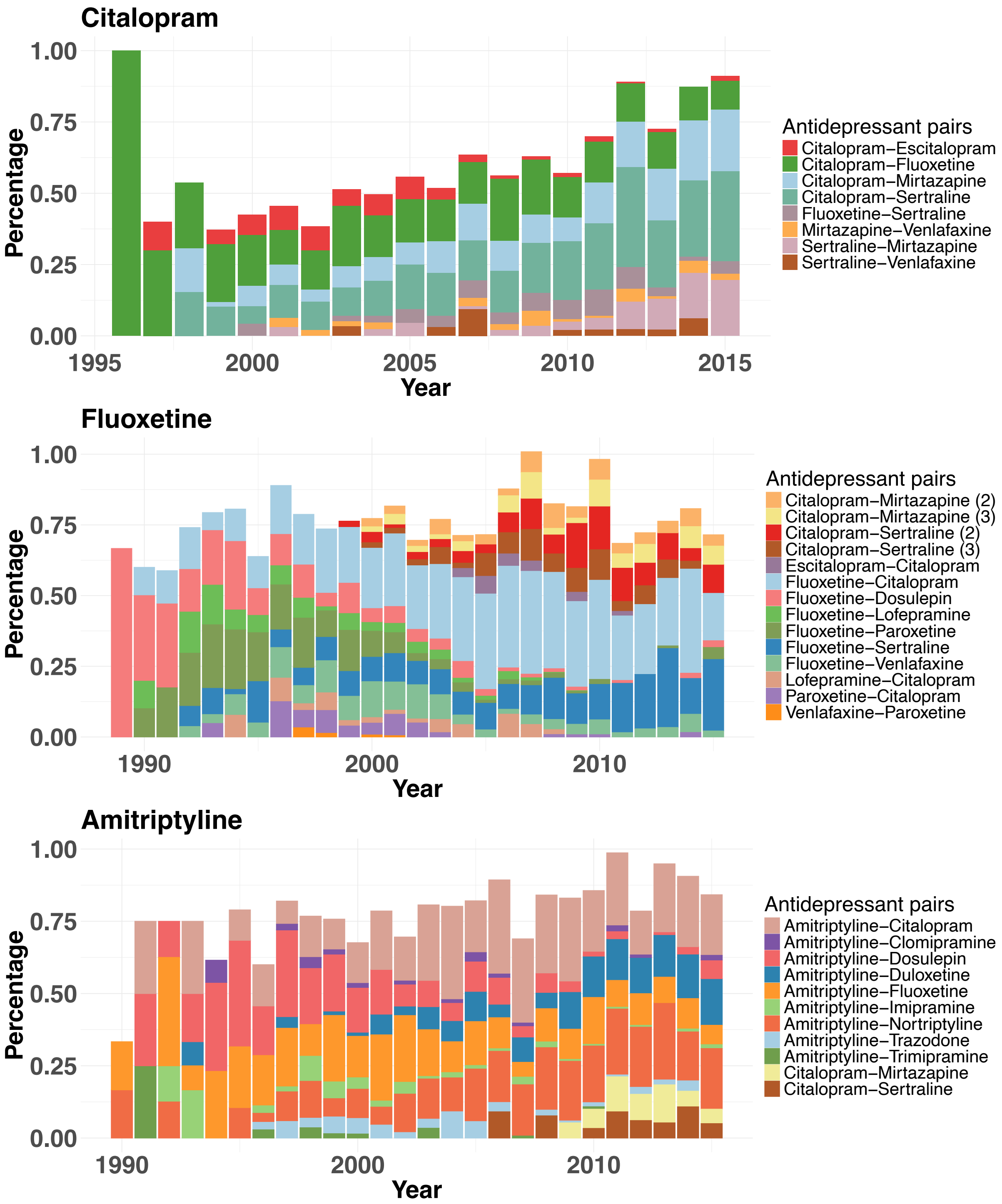


All antidepressant pairs shown in the figure reached statistical significance after multiple corrections. In fluoxetine, the number after the drug pair of citalopram-mirtazapine and citalopram-sertraline showed the order of the drug pair appeared in the antidepressant pathways, where ‘2’ indicated the drug pair was the second and third antidepressant, and ‘3’ represented the drug pair was the third and fourth antidepressant.

Supplementary Figure 9. Early and late discontinuation rates at the first ten prescribed antidepressants

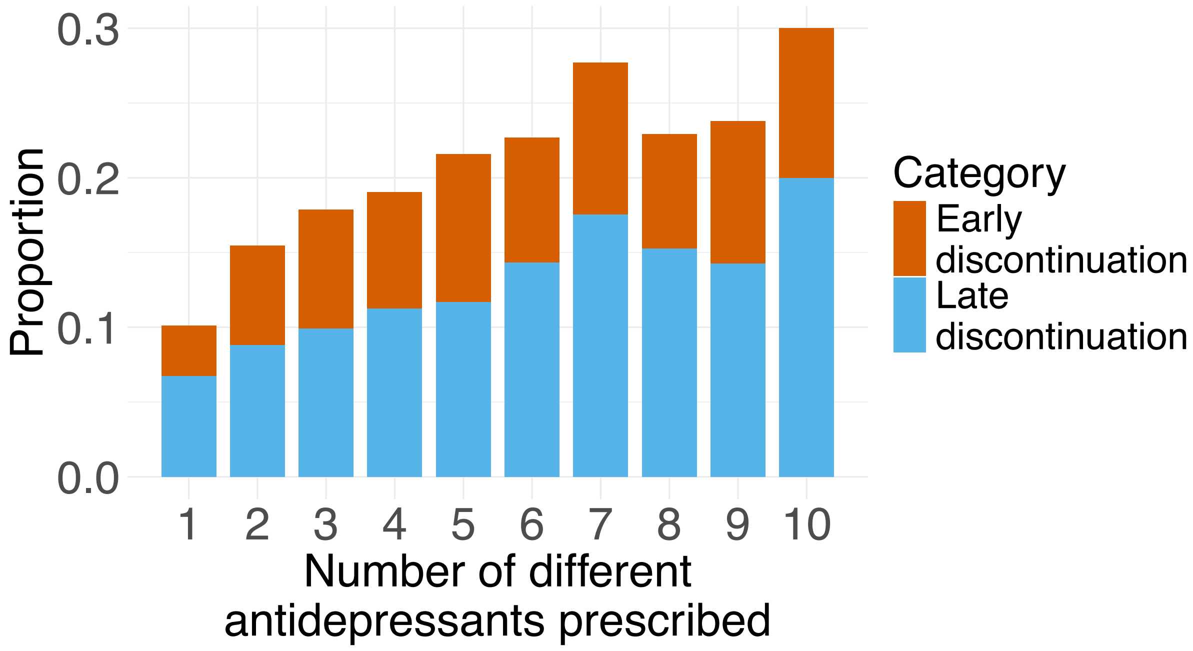

Supplementary Figure 10. Early and late discontinuation rates of each antidepressant class across number of different antidepressants prescribed

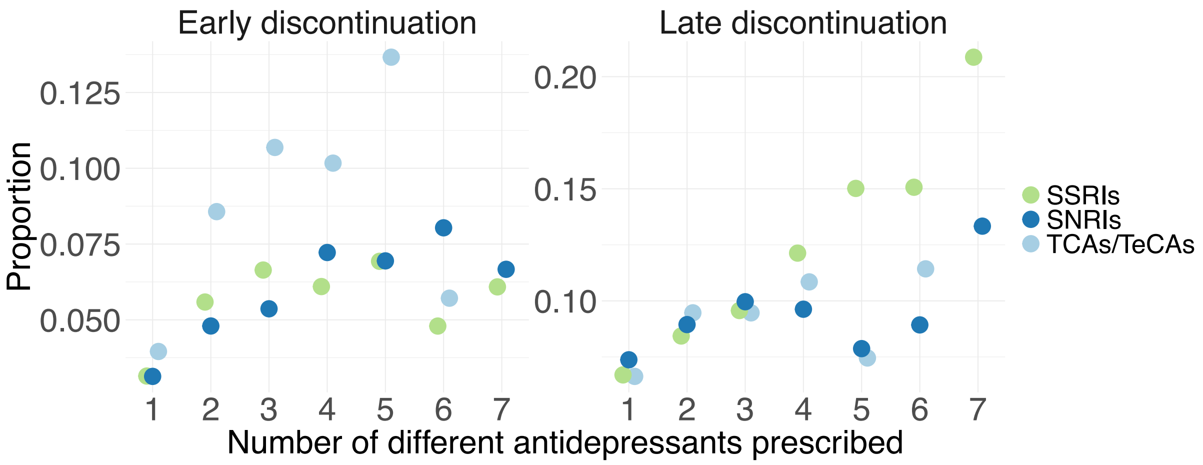


Only antidepressant class with sample size > 50 at each prescription were shown

Supplementary Figure 11. GWAS association of the number of antidepressant changes in all population

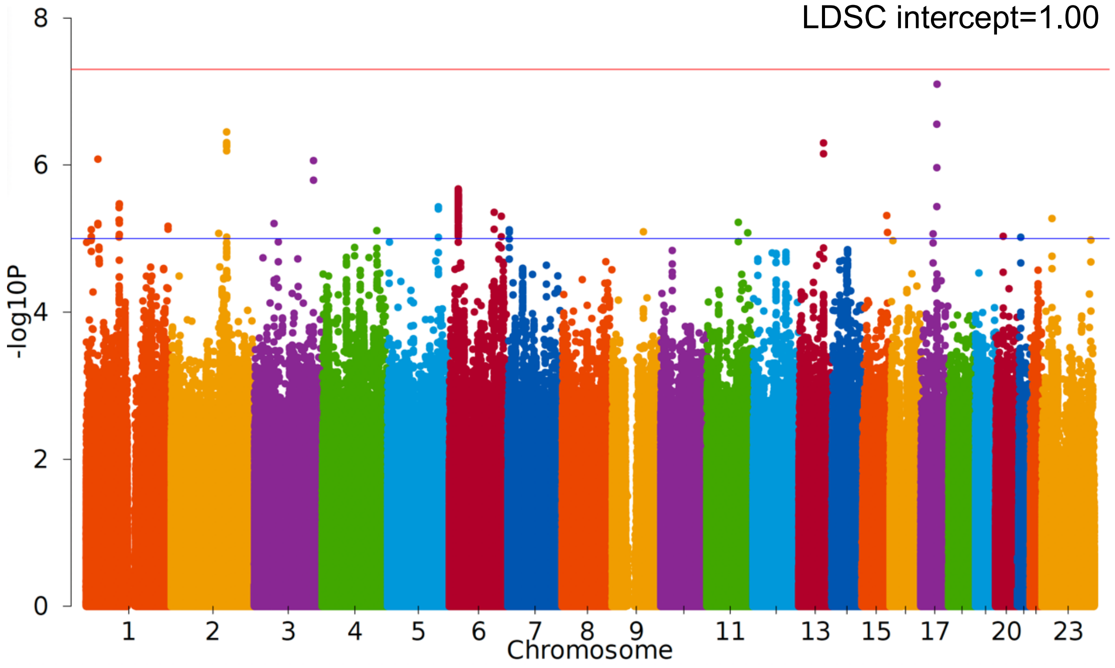


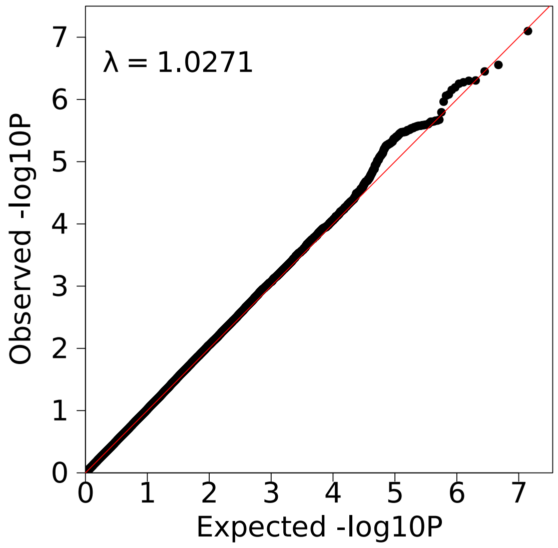

Supplementary Figure 12. GWAS association of the number of antidepressant changes in the depression group

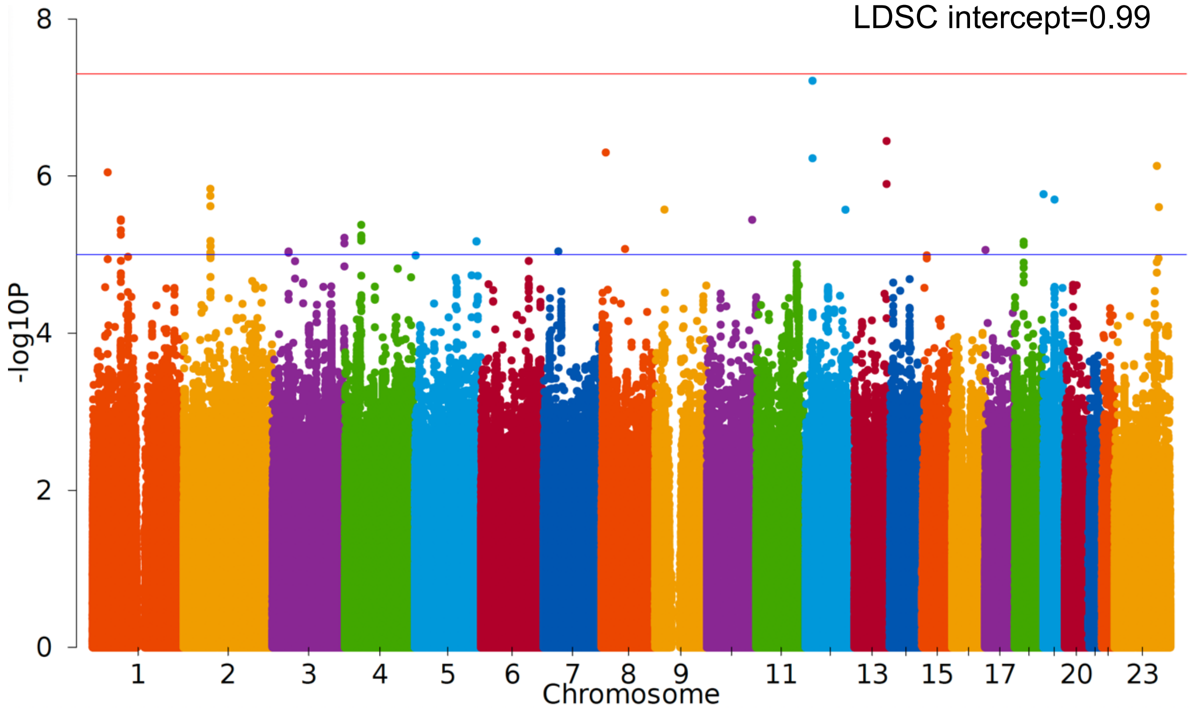


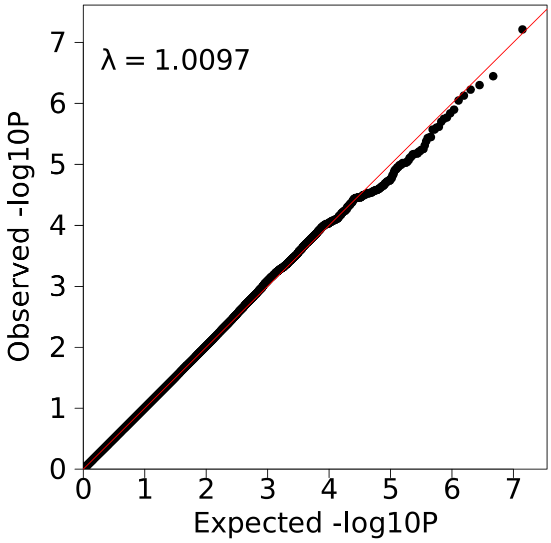

Supplementary Figure 13. GWAS association of the number of antidepressant changes in the non-depression group

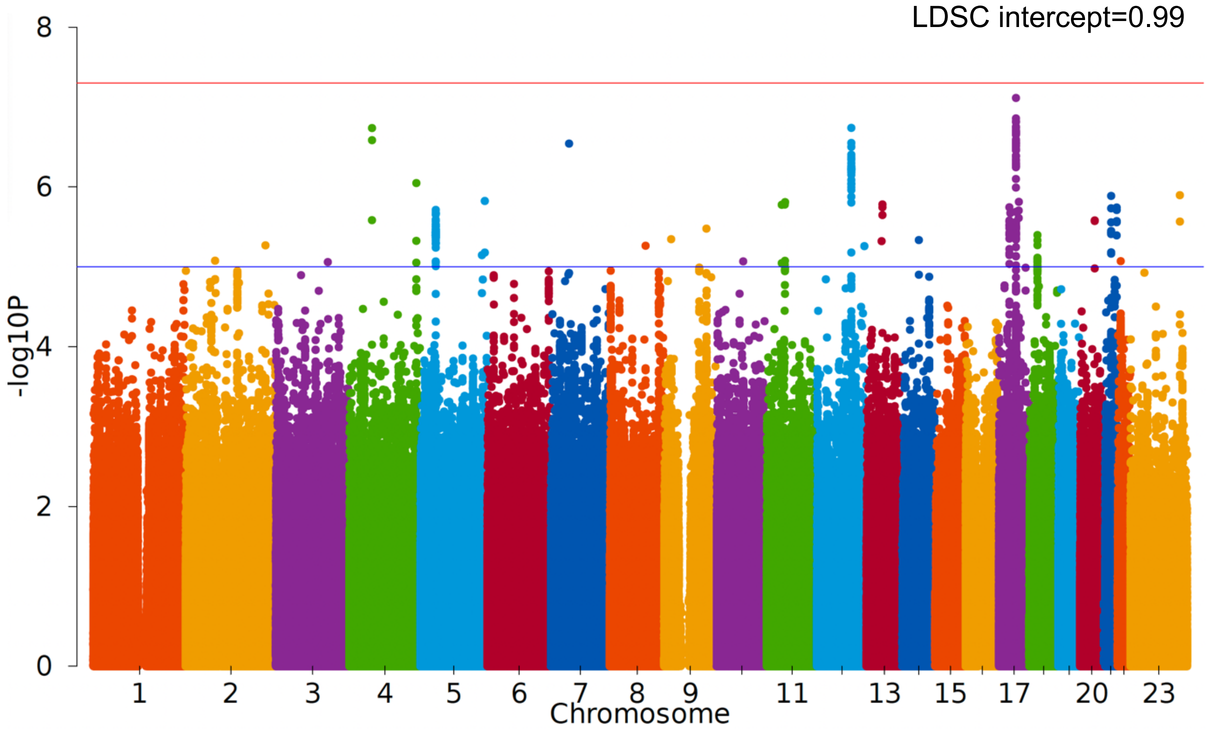


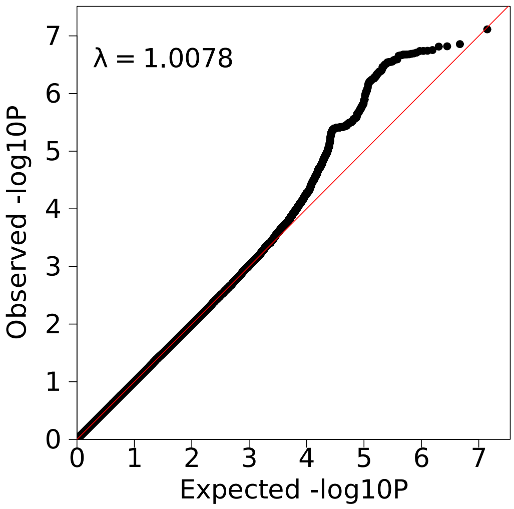

Supplementary Figure 14. GWAS association of SSRI early discontinuation

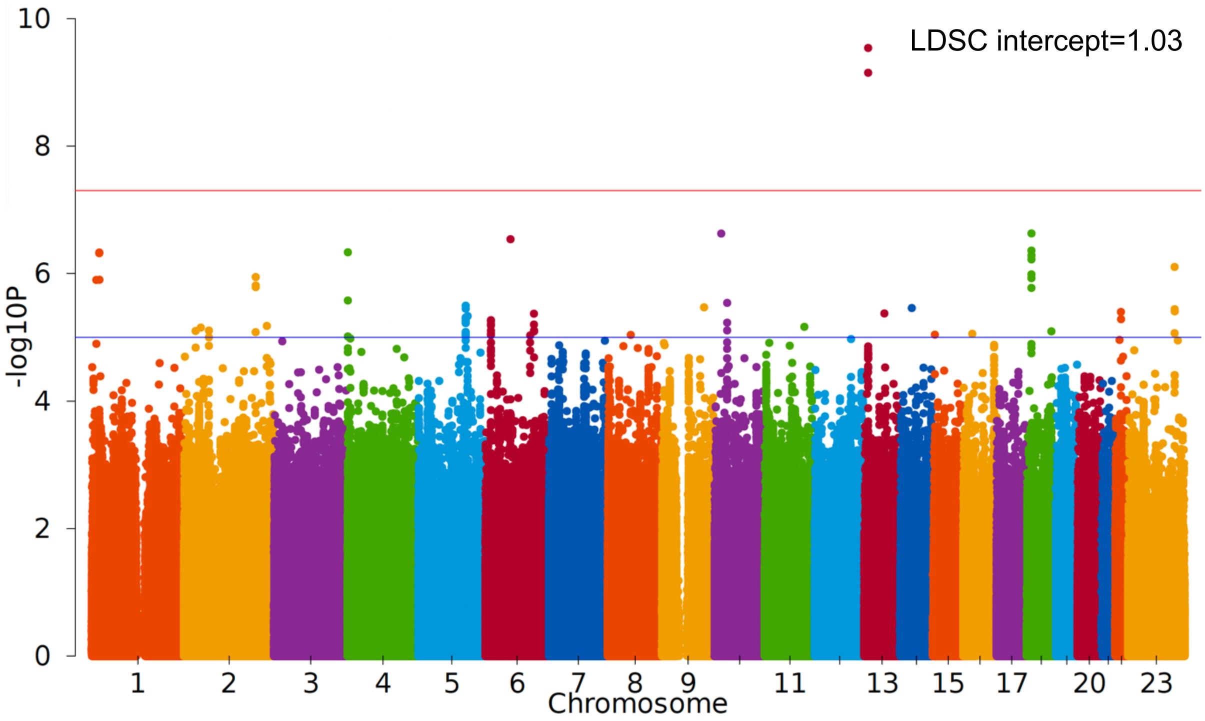


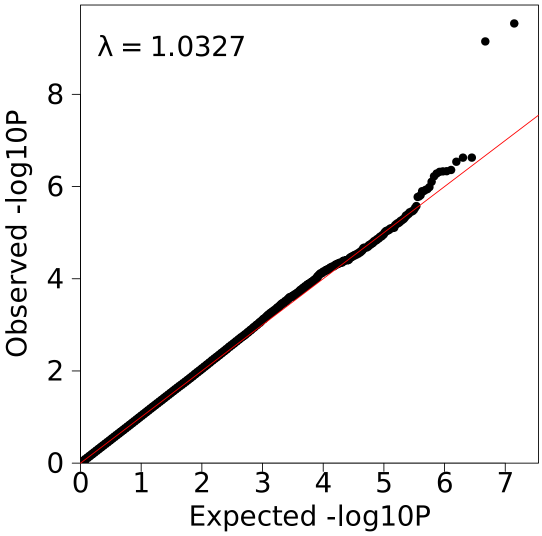

Supplementary Figure 15. GWAS association of SSRI late discontinuation

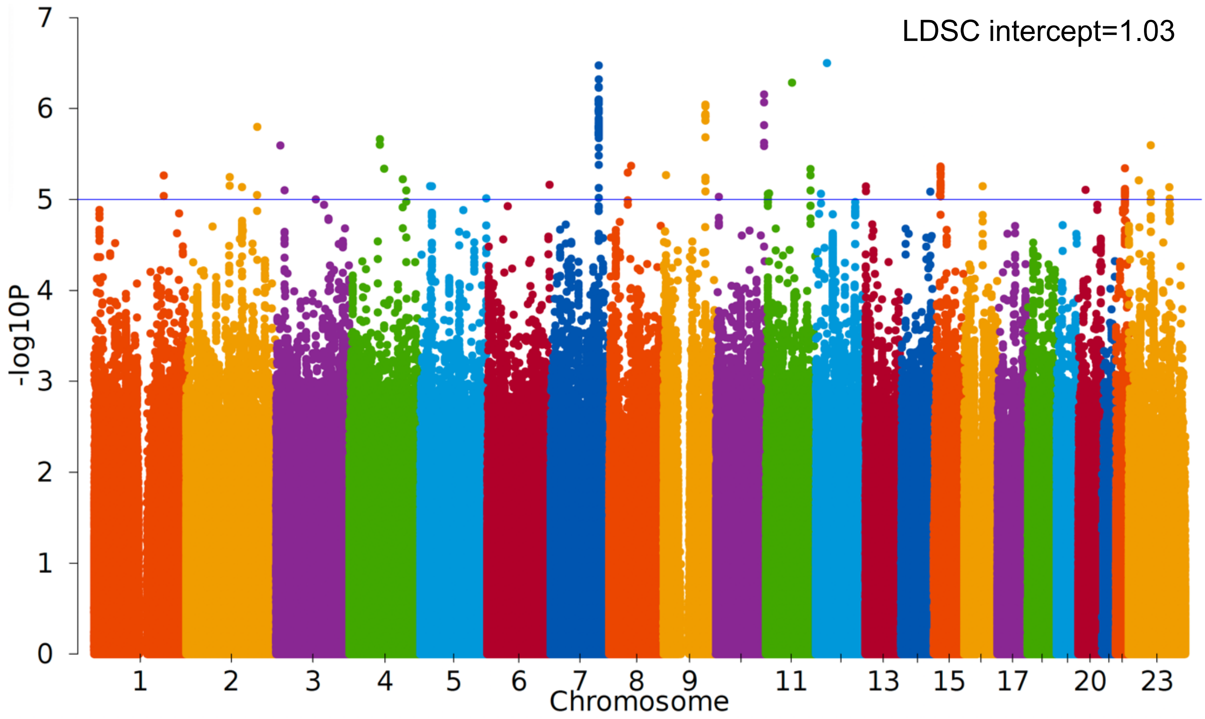


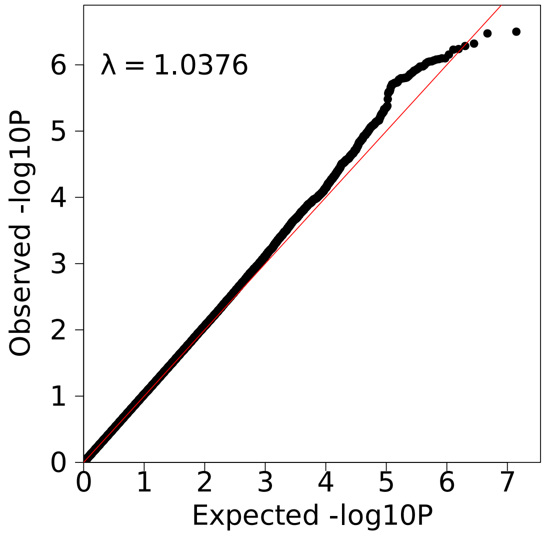

Supplementary Figure 16. SNP heritability (SNP-h2) estimated by GCTA and LDSC

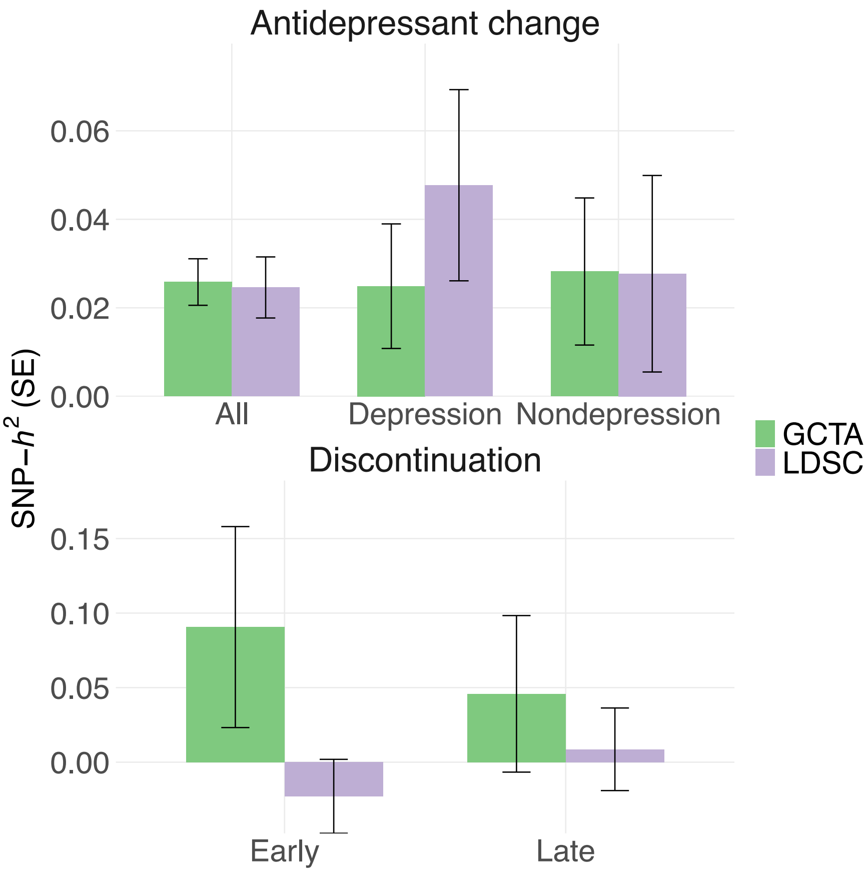


Each error bar showed standard error of the heritability estimate.

#### Supplementary Tables

Supplementary Table 1. Prescribed antidepressants in the UK Biobank

| Drug classes | Antidepressants |
|---|---|
| SSRIs | citalopram, dapoxetine, escitalopram, fluoxetine, fluvoxamine, paroxetine, sertraline |
| SNRIs | duloxetine, venlafaxine |
| SARIs | nefazodone, trazodone |
| TCAs | amitriptyline, amoxapine, clomipramine, dosulepin, doxepin, imipramine, lofepramine, nortriptyline, protriptyline, trimipramine |
| TeCAs | maprotiline, mianserin, mirtazapine |
| Others | agomelatine, amitriptyline_perphenazine, bupropion, flupentixol, isocarboxazid, moclobemide, nortriptyline_fluphenazine, phenelzine, reboxetine, tranylcypromine, tranylcypromine_trifluoperazine, viloxazine, vortioxetine |

Supplementary Table 2. Characteristics of individuals with antidepressant prescription in depression and non-depression groups.

| | Depression | | Non-depression | |
|---|---|---|---|---|
| | N of individuals | % | N of individuals | % |
| Sex | | | | |
| Female | 19302 | 68.1 | 14363 | 58.5 |
| Male | 9030 | 31.9 | 10180 | 41.5 |
| Median age at first prescription | 52 | - | 58 | - |
| Median follow-up time (years) | 12.1 | - | 7.9 | - |

Supplementary Table 3. The top ten most prescribed antidepressants in the UK Biobank

| | Depression | | Non-depression | |
|---|---|---|---|---|
| | N of individuals | % in the depression group | N of individuals | % in the non-depression group |
| SSRIs | | | | |
| Citalopram | 13228 | 46.7 | 2734 | 11.1 |
| Fluoxetine | 11335 | 40.0 | 2039 | 8.3 |
| Sertraline | 6786 | 24.0 | 1330 | 5.4 |
| Paroxetine | 3835 | 13.5 | 509 | 2.1 |
| TCAs | | | | |
| Amitriptyline | 194 | 0.7 | 18426 | 75.1 |
| Dosulepin | 2275 | 8.0 | 1366 | 5.6 |
| Lofepramine | 1976 | 7.0 | 232 | 0.9 |
| SNRIs | | | | |
| Venlafaxine | 3059 | 10.8 | 382 | 1.6 |
| Others | | | | |
| Mirtazapine | 4038 | 14.3 | 806 | 3.3 |
| Trazodone | 1425 | 5.0 | 360 | 1.5 |

Supplementary Table 5. Early and late discontinuation rates across antidepressant classes in individuals with depression

| | Early discontinuation (side effects) | Late discontinuation (nonresponse) |
|---|---|---|
| Total | 8.8% | 11.8% |
| SSRIs | 5.9% | 9.9% |
| SNRIs | 5.2% | 9.2% |
| TCAs | 7.3% | 8.8% |

Supplementary Table 7. Genetic correlation among antidepressant change and discontinuation phenotypes

| Phenotype 1 | Phenotype 2 | rG | se | P |
|---|---|---|---|---|
| Antidepressant change depression | Antidepressant change non-depression | 0.37 | 0.93 | 0.69 |
| Early discontinuation | Late discontinuation | 0.042 | 0.20 | 0.83 |
| Early discontinuation | Antidepressant change depression | 0.058 | 0.35 | 0.87 |
| Early discontinuation | Antidepressant change non-depression | -0.13 | 0.46 | 0.78 |
| Late discontinuation | Antidepressant change depression | 0.56 | 0.33 | 0.086 |
| Late discontinuation | Antidepressant change non-depression | 0.64 | 0.59 | 0.28 |

#### Supplementary Methods

##### Antidepressant prescription for individuals with or without depression

In the UK Biobank, all primary care prescriptions were classified by a number of coding terms including Read v2, British National Formulary (BNF), and Dictionary of Medicines and Devices (dm+d) [1], collected from four primary care computer system suppliers: England TTP, England Vision, Wales EMIS/Vision, and Scotland EMIS/Vision. Each prescription record was annotated with the corresponding medication chemical name (e.g. citalopram, fluoxetine), and drug class (e.g. Selective serotonin reuptake inhibitors (SSRIs), Tricyclic antidepressants), following the table available at: https://doi.org/10.1101/2020.08.24.20178715. In total, 37 antidepressants were included in this study (Supplementary Table 1). For clinical events or prescription dates that preceded or same with a participant's date/year of birth, or were recorded as future dates, UK Biobank assigned predefined placeholder values (01/01/1901, 02/02/1902, 03/03/1903 and 07/07/2037), and these values were treated as missing in our analyses. More detailed description of the primary care prescription records can be found in Fabbri et al [1].

In this study, individuals prescribed antidepressants with depression were defined as those who had at least one primary care diagnosis of depressive disorders and no diagnoses for any psychotic disorders, bipolar disorder, or substance use disorders in their primary care records [1]. These diagnoses were classified by the clinical coding terms version 2 (Read v2) and version 3 (CTV3 or Read v3), which have been used in UK primary care since 1985. Following the study from Fabbri et al. [1], both Read v2 and Read v3 clinical codes were classified into diagnostic groups such as depressive disorders, bipolar disorders, psychosis, and substance use disorders, and the Read v2 clinical codes were mapped to the corresponding Read v3 clinical codes (https://doi.org/10.1101/2020.08.24.20178715).

For individuals prescribed antidepressants without depression, they were included if they had at least one antidepressant prescription but without any depression measures identified in the UK Biobank. This included:

1) Primary care diagnoses of depressive disorders as described above [1].

2) Broad depression: self-reported help-seeking behaviour for mental health difficulties, defined as Individual answering 'yes' to either of the following questions: 'Have you ever seen a general practitioner (GP) for nerves, anxiety, tension, or depression?' (data field 2090) or 'Have you ever seen a psychiatrist for nerves, anxiety, tension or depression?' (data field 2100) [2].

3) Lifetime depression: defined using the Composite International Diagnostic Interview Short Form (CIDI-SF) that was part of the second Mental Health Questionnaire (MHQ) [3]. Criteria for lifetime major depressive episode were in accordance with the DSM-V.

4) Self-reported depression: past and current depression self-reported to the interviewer (a trained nurse) (data field 20002), or answering 'yes' to the question 'During the past six months have you had depression?' (data field 120044).

5) Hospital inpatient diagnosis: based on 3-digit ICD10 codes for F32 Depressive episode, F33 Recurrent depressive disorder, F34 Persistent mood [affective] disorders, F38 Other mood [affective] disorders, F39 Unspecified mood [affective] disorder, from both primary (data field 41202) and secondary diagnoses (data field 41204) [2].

There were 82,633 individuals who were prescribed at least one recorded antidepressant prescription. Among them, 28,332 individuals had at least one primary care diagnosis of depression, and 24,543 individuals did not have any depression identified from the above measures. The remaining 29,758 individuals did not meet the criteria for either group, so they were excluded for the subgroup analyses.

**Antidepressant discontinuation likely due to intolerable side effects or nonresponse in individuals with depression**

We identified two discontinuation phenotypes of SSRIs in the depression group, where early discontinuation was likely due to intolerable side effects and late discontinuation was likely due to nonresponse. For early discontinuation, controls were defined as individuals prescribed any SSRIs more than six weeks and at least three times in a depression episode to ensure they do not suffer from side effects. For late discontinuation, controls were restricted to individuals prescribed any SSRIs for at least six weeks and more than three times in a depression episode, and they either switched to another antidepressant with a break longer than 90 days or had no switch to another drug afterwards. This is to ensure individuals received adequate antidepressant treatment duration in a depression episode without changing to another antidepressant due to nonresponse. Individuals who experienced any early or late discontinuation of SSRI treatment were excluded from their respective control groups.

**Genotype and imputation**

Genome-wide genotyping of all UK Biobank participants was performed using two overlapping arrays, containing ~800,000 markers. Quality control of autosomal genotype data was conducted to correct for potential confounding factors such as genotyping array effects, batch effects, and deviations from Hardy–Weinberg equilibrium (HWE) [4]. SNPs were excluded if they showed high level of missingness (> 2%) or significant deviation from HWE ($p < 1 \times 10^{-8}$). Individuals were removed if they exhibited abnormal heterozygosity (as defined in centralized QC), excessive genotype missingness (> 5%), third-degree or closer relatedness (KING $r < 0.044$), or discordance between genetic sex and self-reported gender. Population structure was evaluated using principal component analysis (PCA), and individuals of European ancestry were identified using four-means clustering based on the first two genetic principal components. Genotype imputation was performed using the Haplotype

Reference Consortium (HRC) and UK10K reference panels [5]. Quality control was performed on the imputed genotype, restricting analyses to variants with imputation quality INFO score < 0.4, minor allele frequency > 0.01, missing rate < 0.05, and Hardy-Weinberg equilibrium exact test p-value > $1\times10^{-8}$.

**Heritability estimate and genetic correlation**

We applied both Genome-wide Complex Trait Analysis (GCTA) and linkage disequilibrium score regression (LDSC) [6,7] to evaluate SNP-based heritability. For binary phenotypes, heritability estimates were transformed to the liability scale using the prevalence observed in the dataset. GCTA models were adjusted for batch, array, the first ten genetic principal components, data provider, birth year, and year of the first prescription. Genetic correlations with selected psychiatric and physical traits were computed using LDSC, constraining the regression intercept to 1. The FDR method was applied for multiple correction across all genetic correlation tests.

**Polygenic score**

Polygenic scores (PGSs) were calculated using SBayesR, which is a Bayesian regression method to estimate SNP effect sizes while accounting for linkage disequilibrium and polygenic architecture [8]. For continuous phenotypes like antidepressant change, a Gamma regression was applied to assess the explained variance of PRS associations. For binary phenotypes including early and late SSRI discontinuation, logistic regression was applied, with Nagelkerke's R2 used to evaluate the explained variance. The population prevalence was assumed same to the dataset. All models were adjusted for the first ten ancestry-specific principal components, data provider, birth year, year of the first prescription and genotyping batch. FDR correction was applied across all PGS tests.
